# Achieving engagement with a Deliberate Practice programme using portable laparoscopy simulators - “Incentivised Laparoscopy Practice”

**DOI:** 10.1101/2020.11.13.20225839

**Authors:** Kenneth G Walker, Jennifer Cleland, Paul M Brennan, Vivienne I Blackhall, Laura G Nicol, Adarsh Shah, Satheesh Yalamarthi, Mark Vella

## Abstract

**Background:** The transfer validity of portable laparoscopy simulation is well established. However, attempts to integrate take-home simulation into surgical training have met with inconsistent engagement, as reported in our 2014-15 study of an Incentivised Laparoscopy Practice (ILP) programme. Our subsequent multi-centre study examined barriers and facilitators, informing revisions of the programme for 2018-20. We now report engagement with the revised versions.

**Methods:** In ILP v2.1 and 2.2, two consecutive year-groups of new CSTs (n= 48 and 46) were loaned portable simulators. The 6-month programme included induction, technical support, and intermittent feedback. Six tasks were prescribed, with video instruction and charting of metric scores. Video uploads were required and scored by faculty. A pass resulted in an eCertificate, expected at Annual Review. ILP was set within a wider reform, “Improving Surgical Training”.

**Results:** ILP v2.1 and 2.2 saw pass rates of 94% and 76% (45/48 and 35/46 trainees respectively), compared with only 26% (7/27) in v1, despite the v2.1 and v2.2 groups having less electronic gaming experience. In the ILP v2.2 group, 73% reported their engagement was adversely affected by COVID19 redeployments.

**Conclusions:** Simply providing kit, no matter how good, is not enough. To achieve trainee engagement with take- home simulators, as in ILP v2, a whole programme is required, with motivated learning, individual and group practice, intermittent feedback, and clear goals and assessments. ILP is a complex intervention, best understood as a “reform within a reform, within a context.” This may explain why trainee engagement fell away during early pandemic conditions.

**WHAT IS ALREADY KNOWN ON THIS SUBJECT:**

- Attaining automation of motor skills is essential to free up operating surgeons’ attention for higher cognitive functions.
- Laparoscopic operating skills can transfer from simulation to the operating room, and deliberate practice is the most important variable in the development of expertise.
- Simply providing take-home portable simulators to surgical trainees, even with online training programmes, is insufficient to facilitate consistent deliberate practice by more than a minority of trainees.

**WHAT THIS STUDY ADDS:**

- A package of evidence-based reforms transformed participation of Core Surgical trainees in a 6-month programme of practice using take-home portable simulators, resulting in near- 100% engagement.
- Such reforms are complex, including motivators for learning, individual and group practice, intermittent feedback, clear goals and assessments, and adoption into a wider curriculum reform called “Improving Surgical Training”.
- The improved engagement with this form of remote simulation-based training did not continue in the face of a national “lockdown” for the COVID19 pandemic, where there was widespread redeployment of trainees.

## INTRODUCTION

Laparoscopic operating skills gained in the early years of surgical training have been reported to fall short of desirable standards, ^1^ even in contexts where formal assessment of these skills has been introduced to support quality assurance (e.g. the Fundamentals of Laparoscopic Surgery exam ^2^). The reasons for this gap between actual and desired levels of competence are complex but may be related, at least to some extent, to trainee/resident practice using simulation being mostly *ad hoc* and self-directed, rather than deliberate and structured.

Drawing on the principles and outcomes reported widely in disciplines such as music and sports, ^3^ some training programmes in the UK and Ireland have provided trainees with resources and guidance for deliberate Practice (DP) in their own time using portable simulators and a modular curriculum. Evidence suggests that DP is the most important variable in the development of expertise ^3–5^ as only once automation is achieved is a trainee’s attention (“bandwidth”) freed up for the higher cognitive functions of surgery, i.e., non-technical skills. ^6,7^ However, in contrast with professional musicians and sports people, it has proved surprisingly difficult to achieve consistent engagement with DP amongst surgeons in training. ^8,9^ Yet if trainees do not engage with DP, then they may struggle to gain necessary skills within limited training hours and restricted access to patients.

In the first of our group’s two previous studies, ^10^ we reported that only 7 of 21 Scottish Core Surgical trainees (residency year 1 and 2) completed a modular DP programme using take-home laparoscopic simulators (“Incentivised Laparoscopy Practice” (ILP)) in 2014-15. This lack of engagement was noted despite the robust and portable chosen simulators (eoSim, eoSurgical, Edinburgh, UK; eosurgical.com) being loaned to the trainees free-of-charge. The simulator’s instrument tracking software complemented the well-validated online curriculum of modular tasks and instructional videos. Cumulatively, the hardware and software enabled trainees to see their metric scores and upload videos for structured scoring by faculty (app.surgtrac.com).^11^ Moreover, trainees completing and passing the programme were rewarded with an incentive (an eCertificate) to provide evidence to their trainers. However, the reasons given for non-completion included technical problems, competing demands on time (e.g., College examinations [MRCS]), career intentions in non-abdominal subspecialties, and lack of understanding of the educational rationale behind the programme.

Having noted similar experiences reported by colleagues in the Republic of Ireland and in the Wessex and Severn deaneries of England, we proceeded to a second study in 2017. ^12^ Its design was also informed by a thematic analysis of 22 papers published in 2016 identifying the core factors for successful “off-site training of laparoscopic skills”. ^13^ The aim of this second study was to further explore the barriers and facilitators to engagement with ILP in all 4 geographical regions via focus groups with trainees and trainers, as a basis for redesign of our ILP programme in Scotland.

Now in this third paper, we report how these principles and the information gathered from our own studies, informed the Scottish Surgical Simulation Collaborative’s (SSSC) further development of a DP programme - ILP - and affected trainee engagement.

## METHODS

As this was a service quality improvement process, ethics approval was not required.

The focus group findings from the second study ^12^ and how we responded to these in the redesign process are presented in Table 1. Note that some of these changes were incorporated for the Scottish Core Surgical year 1 trainees of 2018-2019 (n=48) (“ILP 2.1”). For the 46 trainees in year 2019-20, we made some further refinements for (“ILP 2.2”), to embed it even further into the wider core surgical training system. These were: shifting the timing of induction and assessments, group practice in Boot Camp, and a requirement to upload six videos (one for each module) for scoring by one of a panel of trainers using a structured assessment proforma similar to OSATS (Objective Structured Assessment of Technical Skill ^14^, figure 2). The pass mark (an average 9/21 per video) was determined by previous years’ data, being set just below the typical level that was achieved only by trainees who had put in significant hours of practice.

**Table 1:**
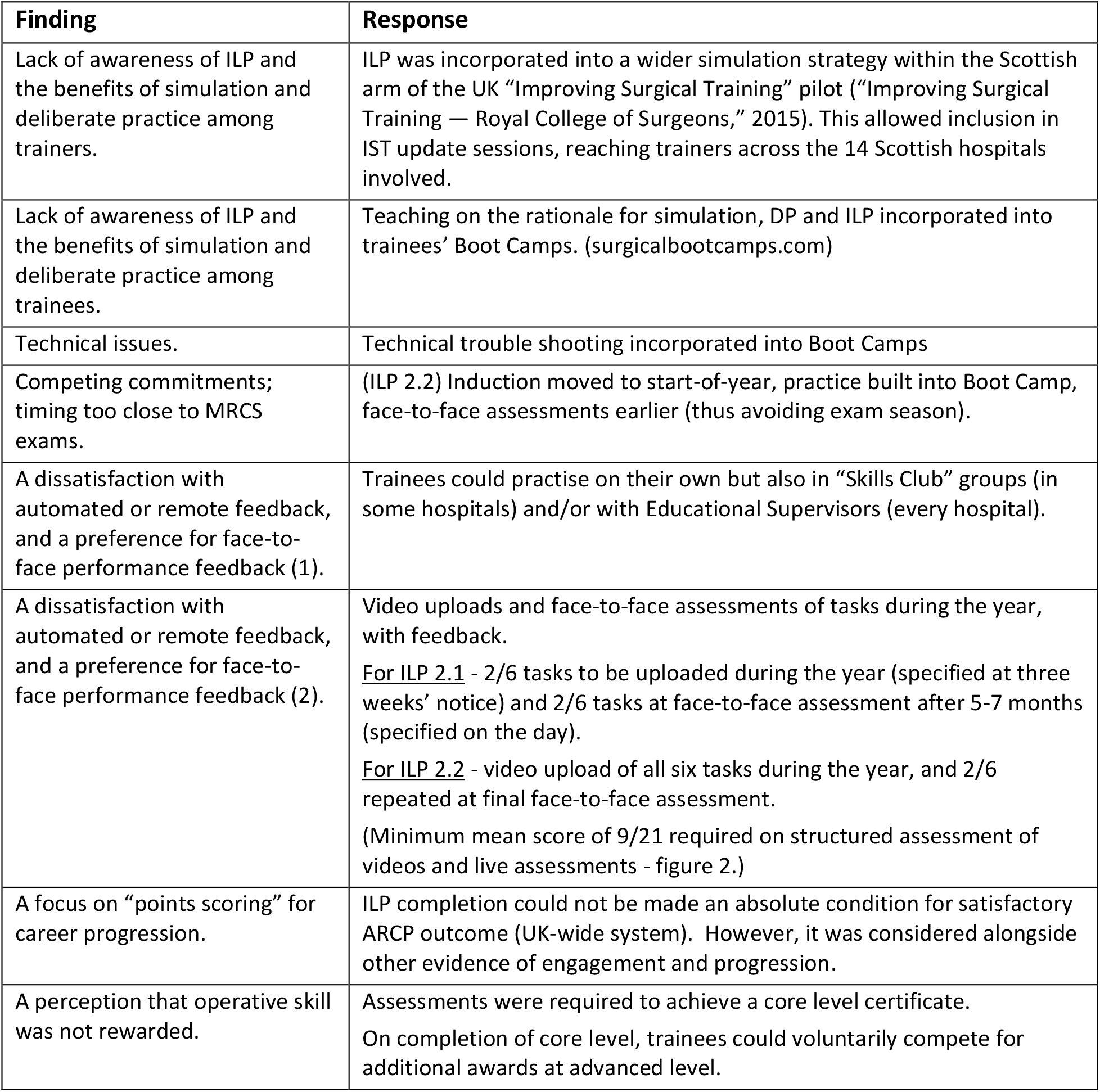
ILPS 2: evidence-informed curriculum reform.

| **Finding** | **Response** |
| --- | --- |
| Lack of awareness of ILP and the benefits of simulation and deliberate practice among trainers. | ILP was incorporated into a wider simulation strategy within the Scottish arm of the UK “Improving Surgical Training” pilot (“Improving Surgical Training — Royal College of Surgeons,” 2015). This allowed inclusion in IST update sessions, reaching trainers across the 14 Scottish hospitals involved. |
| Lack of awareness of ILP and the benefits of simulation and deliberate practice among trainees. | Teaching on the rationale for simulation, DP and ILP incorporated into trainees’ Boot Camps. (surgicalbootcamps.com) |
| Technical issues. | Technical trouble shooting incorporated into Boot Camps |
| Competing commitments; timing too close to MRCS exams. | (ILP 2.2) Induction moved to start-of-year, practice built into Boot Camp, face-to-face assessments earlier (thus avoiding exam season). |
| A dissatisfaction with automated or remote feedback, and a preference for face-to-face performance feedback (1). | Trainees could practise on their own but also in “Skills Club” groups (in some hospitals) and/or with Educational Supervisors (every hospital). |
| A dissatisfaction with automated or remote feedback, and a preference for face-to-face performance feedback (2). | Video uploads and face-to-face assessments of tasks during the year, with feedback.<br><u>For ILP 2.1</u> - 2/6 tasks to be uploaded during the year (specified at three weeks’ notice) and 2/6 tasks at face-to-face assessment after 5-7 months (specified on the day).<br><u>For ILP 2.2</u> - video upload of all six tasks during the year, and 2/6 repeated at final face-to-face assessment.<br>(Minimum mean score of 9/21 required on structured assessment of videos and live assessments - figure 2.) |
| A focus on “points scoring” for career progression. | ILP completion could not be made an absolute condition for satisfactory ARCP outcome (UK-wide system). However, it was considered alongside other evidence of engagement and progression. |
| A perception that operative skill was not rewarded. | Assessments were required to achieve a core level certificate.<br>On completion of core level, trainees could voluntarily compete for additional awards at advanced level. |

**Figure 1.**
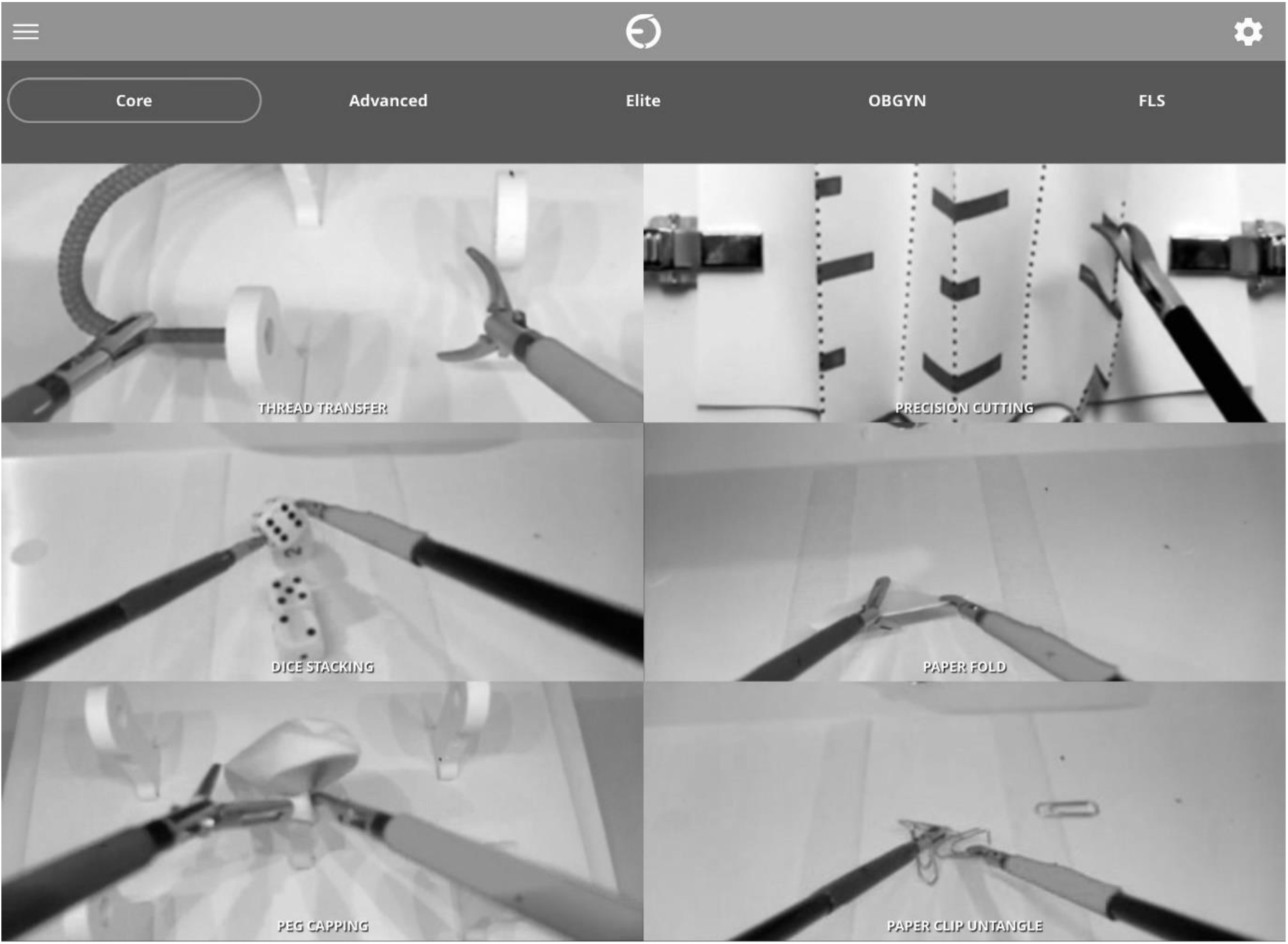
The 6 modular tasks comprising the core level programme (screenshot from eosurgical.com).

**Figure 2.**
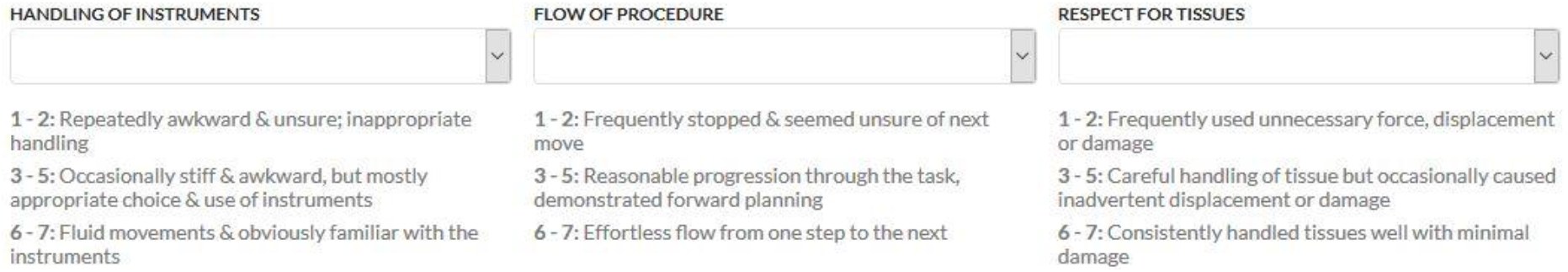
Web-based structured assessment form for scoring uploaded videos (app.surgtrac.com).

After completion of Year 1 of ILP/Core Surgical Training, each year group was sent an e-survey. This requested demographic information plus previous laparoscopy and gaming experience, so groups that could be directly compared with the ILP 1 group ^10^.

## RESULTS

The ILP 1 study in 2014-15 had recruited only about a quarter of the Scottish CST years 1 and 2 cohorts to the project on a voluntary basis (27 trainees). In contrast, once the revised programme was made an expected part of the IST simulation strategy from 2018, the ILP 2.1 and 2.2 cohorts then included the entire 1^st^ year group each year: 48 trainees in 2018-19 and 46 trainees in 2019-20.

Table 2 shows the proportion of each cohort completing the programme and attaining the OSATS pass mark of at least 9/21 (whether at final face-to-face assessment in ILP 1 and 2.1, or across 6 uploaded videos in ILP 2.2). This completion rate rose from just 26% in ILP 1 to 94% and 76% in ILP 2.1 and 2.2 respectively. In the latter group, of the 22 who completed the survey, the majority (16 trainees) said their engagement had been adversely affected by the COVID19 pandemic, which affected Scotland mostly from March 2021, i.e. about 2 months before the closing date for video uploads.

**Table 2.** Comparison of the ILP 1, 2.1 and 2.2 groups.

|  |  | ILP 1<br>(2014-15) | ILP 2.1<br>(2018-19) | ILP 2.2<br>(2019-20) |
| --- | --- | --- | --- | --- |
| **Part of IST pilot** |  | No | Yes | Yes |
| **No of trainees participating** |  | 27 | 48 | 46 |
| **Core level result** | **Completed & passed** | 7 (26%) | 45 (94%) | 35 (76%) |
|  | **Failed to complete & pass** | 20 (74%) | 2 (4%) | 11 (24%) |
|  | **Absent from final assess<sup>t</sup>** | 0 | 1 (2%) |  |
| **No responding to survey** |  | 23/27 | 25/48 | 22/46 |
| **Engagement adversely affected by pandemic?** | **Yes** |  |  | 16 (73%) |
|  | **No** |  |  | 4 (18%) |
|  | **No answer** |  |  | 2 (9%) |
| **Age** | **24-29** | 19 (83%) | 12 (48%) | 15 (68%) |
|  | **30-35** | 4 (17%) | 10 (40%) | 7 (32%) |
|  | **>35** | 0 | 3 (12%) | 0 |
| **Sex** | **Male** | 17 (63%) | 12 (48%) | 11 (50%) |
|  | **Female** | 10 (37%) | 11 (44%) | 10 (46%) |
|  | **Prefer not to say / answer** | 0 | 2 (8%) | 1 (5%) |
| **Prior laparoscopic surgery experience** | **Assisted lap surg 1-5 x** |  | 4 (16%) | 6 (27%) |
|  | **Assisted lap surg >5 x** |  | 14 (56%) | 10 (45%) |
|  | **Mobilised GB or appendix** |  | 11 (44%) | 9 (41%) |
|  | **1-5 prior lap sim sessions** |  | 6 (24%) | 5 (23%) |
|  | **>5 prior lap sim sessions** |  | 1 (4%) | 0 |
| **Gaming experience** | **>3 hrs /week** | 7 (31)% | 3 (12%) | 2 (9%) |
|  | **None or < 3hrs /week** | 15 (65%) | 22 (88%) | 19 (86%) |
|  | **Did not answer** | 1 (4%) | 0 | 1 (5%) |

The ILP 2.1 and 2.2 trainee cohorts were slightly older (40% and 32% over 30, compared with 17% of the ILP 1 group) and more evenly balanced for sex (48% and 50% male, compared with 63% in ILP 1).

The majority of ILP 2.1 and 2.2 trainees had assisted at laparoscopic surgery before starting the programme; 44% and 42% had previously mobilised a gallbladder or an appendix (see table 2). No data on prior laparoscopic experience were available from the earlier ILP 1 group.

Whereas 31% of the earlier ILP 1 group had reported prior electronic gaming experience of at least 3 hours a week, only 12% and 9% of the ILP 2.1 and 2.2 groups did so.

## DISCUSSION

To address a gap in knowledge, we completed a series of studies examining how best to engage core surgical trainees with home-based Deliberate Practice (DP) of laparoscopic skills using box simulators (ILP). We report on how the findings from the initial study were the basis for redesigning ILP, and the redesigned ILP was the first programme to show success with take-home laparoscopic practice in respect of trainee completion and pass rates.

In retrospect, what we designed was a complex educational intervention. In health services research, complex interventions are generally defined as those that involve more than one component. ^15^ The Medical Research Council (MRC) for the development, implementation and evaluation of complex interventions describes several aspects of complexity. ^15,16^ These can be extrapolated to educational interventions such as the one we report ^17^: the number of different components within the intervention (e.g., the different tasks, need to upload tasks, eCertificate); engaging with the task (involving motivation, time management, understanding why DP is relevant to them); who is involved (not only trainees but also educational supervisors); the outcomes (were the gains clear to trainees?); and how tailored the intervention is to individual learners (in respect of, for example, pace, but also wider issues such as proximity to MRCS). All these components interact in non-linear ways which affect how an educational innovation is perceived and engaged with, and ultimately the outcomes it achieves. Simply handing out kit, no matter how good, is not enough. A whole programme is required, with motivated learning, easy, distributed access to practice, intermittent feedback, and clear goals and testing.

We drew heavily on the evidence base for DP and simulation, and our own knowledge of surgical training. Our initial ILP can be considered a feasibility study, to test our procedures, assess engagement/recruitment and retention and determine what worked and what didn’t. In this way, following the cyclical, interactive process proposed by the MRC framework, the findings from that initial study resulted in the thoughtful application of evidence-based frameworks and concepts to the current ILP programme: the introduction of a dedicated taught cognitive component at a Surgical Bootcamp prior to issue of the take-home simulators, ^18,19^ automatic recruitment of all core trainees into the programme, ^20^ incorporating distributed practice with regular formative assessments, ^21^ proficiency-based partial-task training ^22,23^ and faculty engagement. We also addressed technical issues. All changes had the aim of incentivising DP.

It was not only the novel intervention at study that was complex; it in turn took place within an existing complex intervention (surgical training; ^24^) which itself was changing at the time of our studies (e.g. the shift to Improving Surgical Training [IST]). This wider curricular change may have impacted positively on ILP outcomes: the IST proposal recommends simulation as part of the core curriculum, and this is likely to have helped trainee and trainer engagement. ^25,26^ The advent of IST was temporally correlated with an increasing interest in the CST programme, as evidenced by the competition ratios (number of applicants for each successful appointment): 2016 - 2.53, 2017 - 2.56, then for IST 2018 - 2.94, and 2019 - 2.93.

Finally, the context of surgical training is itself not free from other external influences: for example, the COVID19 lockdown (including redeployment of some surgical trainees to other departments) clearly impacted on the engagement and outcomes of the ILP 2.2 group, even though ILP might have been seen by many as the ideal remote training modality for the moment. ^27,28^ This is consistent with our argument that ILP does not work in isolation. It will be interesting to observe engagement in the coming year, once the training programmes have adapted to supporting trainees during the ongoing pandemic.

The multi-faceted nature of ILP and the relatively small number of Core Surgical trainees in Scotland limits evaluation of outcomes. However, the proportion of trainees who completed the core level of ILP in the second group was substantially greater than those who took part in the first intervention. While acknowledging this might have been a cohort effect, we suggest it does indicate that the lessons learned, and changes made after the first ILP improved the effectiveness of ILP. However, was the improved engagement a consequence of one, or a combination, of the changes outlined in Table 1? We have no way of knowing – teasing out the active ingredients of ILP and indeed other surgical education interventions requires further study.

We aim now to apply the lessons to another take-home practice scheme for year 2 of CST, involving vascular anastomoses practised using 3D-printed hydrogel models and kits posted to trainees. We know it will not be enough simply to hand out the kit.

## Data Availability

All related data not included in the manuscript may be made available later on reasonable request.

## Acknowledgements

The following contributed to the Incentivised Laparoscopy Practice (ILP) scoring and feedback panel:- Vivienne Blackhall, Mark Duxbury, Alistair Geraghty, Simon Gibson, Joanna Gray, Morag Hogg, Graham MacKay, Alastair Moses, Laura Nicol, Grenville Oades, Raymond Oliphant, Anna Paisley, Roland Partridge, Benjamin Perakath, Andrew Renwick, Mark Vella, Kenneth Walker, Satheesh Yalamarthi.

The ILP 2.1 and 2.2 programmes of 2018-2021 were part of the simulation strategy delivered within the Scottish part of the UK-wide Improving Surgical Training (IST) pilot, funded by Scottish Government through NHS Education for Scotland (NES).

Support from EOSurgical in providing and maintaining the hardware and software, was provided by Roland Partridge, Paul Brennan and Razvan Ilin.

The administration of ILP 1 was undertaken by the authors Laura Nicol and Kenneth Walker, and of ILP 2.1 and 2.2 by Karen Willey and Lynn Hardie, project officers at the Clinical Skills Managed Educational Network, NES, along with Kenneth Walker.

## REFERENCES

1. Mattar SG, Alseidi AA, Jones DB, et al. General Surgery Residency Inadequately Prepares Trainees for Fellowship. Ann Surg. 2013;258(3):440–449. doi:10.1097/sla.0b013e3182a191ca

2. Peters JH, Fried GM, Swanstrom LL, et al. Development and validation of a comprehensive program of education and assessment of the basic fundamentals of laparoscopic surgery. Surgery. 2004;135(1):21–27. doi:10.1016/S0039-6060(03)00156-9

3. Ericsson KA. Deliberate practice and acquisition of expert performance: a general overview. Acad Emerg Med. 2008;15(11):988–994. doi:10.1111/j.1553-2712.2008.00227.x

4. Fitts P, Posner M. Human Performance. Belmont, CA: Brooks/Cole; 1969.

5. Hamdorf JM, Hall JC. Acquiring surgical skills. Br J Surg. 2000. doi:10.1046/j.1365-2168.2000.01327.x

6. Yule S, Flin R, Paterson-Brown S, Maran N, Rowley D. Development of a rating system for surgeons’ non-technical skills. Med Educ. 2006;40(11):1098–1104. doi:10.1111/j.1365-2929.2006.02610.x

7. Yule S, Flin R, Maran N, Rowley D, Youngson G, Paterson-Brown S. Surgeons’ non-technical skills in the operating room: reliability testing of the NOTSS behavior rating system. World J Surg. 2008;32(4):548–556. doi:10.1007/s00268-007-9320-z

8. Gostlow H, Marlow N, Babidge W, Maddern G. Systematic Review of Voluntary Participation in Simulation-Based Laparoscopic Skills Training: Motivators and Barriers for Surgical Trainee Attendance. J Surg Educ. 2017. doi:10.1016/j.jsurg.2016.10.007

9. Van Dongen KW, Van Der Wal WA, Rinkes IHMB, Schijven MP, Broeders IAMJ. Virtual reality training for endoscopic surgery: Voluntary or obligatory? Surg Endosc Other Interv Tech. 2008. doi:10.1007/s00464-007-9456-9

10. Nicol LG, Walker KG, Cleland J, Partridge R, Moug SJ. Incentivising practice with take-home laparoscopic simulators in two UK Core Surgical Training programmes. BMJ Simul Technol Enhanc Learn. 2016;2(4):112–117. doi:10.1136/bmjstel-2016-000117

11. Hennessey IAM, Hewett P. Construct, concurrent, and content validity of the eoSim laparoscopic simulator. J Laparoendosc Adv Surg Tech A. 2013;23(10):855–860. doi:10.1089/lap.2013.0229

12. Blackhall VI, Cleland J, Wilson P, Moug SJ, Walker KG. Barriers and facilitators to deliberate practice using take-home laparoscopic simulators. Surg Endosc. 2019;33(9):2951–2959. doi:10.1007/s00464-018-6599-9

13. Thinggaard E, Kleif J, Bjerrum F, et al. Off-site training of laparoscopic skills, a scoping review using a thematic analysis. Surg Endosc. 2016;30(11):4733–4741. doi:10.1007/s00464-016-4834-9

14. Martin JA, Regehr G, Reznick R, et al. Objective structured assessment of technical skill (OSATS) for surgical residents. Br J Surg. 1997. doi:10.1002/bjs.1800840237

15. Campbell M, Fitzpatrick R, Haines A, et al. Framework for design and evaluation of complex interventions to improve health. Br Med J. 2000;321(7262):694–696. doi:10.1136/bmj.321.7262.694

16. Craig P, Dieppe P, Macintyre S, Mitchie S, Nazareth I, Petticrew M. Developing and evaluating complex interventions: The new Medical Research Council guidance. BMJ. 2008;337(7676):979–983. doi:10.1136/bmj.a1655

17. Mattick K, Barnes R, Dieppe P. Medical education: A particularly complex intervention to research. Adv Heal Sci Educ. 2013;18(4):769–778. doi:10.1007/s10459-012-9415-7

18. McClusky DA, Smith CD. Design and development of a surgical skills simulation curriculum. World J Surg. 2008;32(2):171–181. doi:10.1007/s00268-007-9331-9

19. Stefanidis D, Heniford BT. The formula for a successful laparoscopic skills curriculum. Arch Surg. 2009;144(1):77–82. doi:10.1001/archsurg.2008.528

20. Chang L, Petros J, Hess DT, Rotondi C, Babineau TJ. Integrating simulation into a surgical residency program: Is voluntary participation effective? Surg Endosc Other Interv Tech. 2007;21(3):418–421. doi:10.1007/s00464-006-9051-5

21. Moulton CAE, Dubrowski A, MacRae H, Graham B, Grober E, Reznick R. Teaching surgical skills: What kind of practice makes perfect? A randomized, controlled trial. Ann Surg. 2006;244(3):400–407. doi:10.1097/01.sla.0000234808.85789.6a

22. Kolozsvari NO, Feldman LS, Vassiliou MC, Demyttenaere S, Hoover ML. Sim one, do one, teach one: Considerations in designing training curricula for surgical simulation. J Surg Educ. 2011;68(5):421–427. doi:10.1016/j.jsurg.2011.03.010

23. Motola I, Devine LA, Chung HS, Sullivan JE, Issenberg SB. Simulation in healthcare education: A best evidence practical guide. AMEE Guide No. 82. Med Teach. 2013;35(10). doi:10.3109/0142159X.2013.818632

24. Cleland J, Walker KG, Gale M, Nicol LG. Simulation-based education: understanding the socio-cultural complexity of a surgical training “boot camp”. Med Educ. 2016;50(8):829–841. doi:10.1111/medu.13064

25. Satava RM. Surgical education and surgical simulation. World J Surg. 2001;25(11):1484–1489. doi:10.1007/s00268-001-0134-0

26. Issenberg SB, McGaghie WC, Petrusa ER, Gordon DL, Scalese RJ. Features and uses of high-fidelity medical simulations that lead to effective learning: A BEME systematic review. Med Teach. 2005;27(1):10–28. doi:10.1080/01421590500046924

27. Okland TS, Pepper JP, Valdez TA. How do we teach surgical residents in the COVID-19 era? J Surg Educ. 2020;77(5):1005. doi:10.1016/j.jsurg.2020.05.030

28. Shah AP, Falconer R, Watson AJM, Walker KG. Teaching surgical residents in the COVID-19 era: The value of a simulation strategy. J Surg Educ. September 2020. doi:10.1016/j.jsurg.2020.08.043

